# Informing an Intervention to Improve Access to Community Pharmacist-Provided Injectable Naltrexone for Formerly Incarcerated Individuals in Wisconsin

**DOI:** 10.1101/2024.09.23.24314214

**Authors:** Jason S. Chladek, Michelle A. Chui

## Abstract

In Wisconsin, opioid use disorder (OUD) is highly prevalent among individuals impacted by the criminal justice system. Medications for opioid use disorder (MOUD), including injectable naltrexone, are crucial for treating OUD and especially important for individuals transitioning out of correctional facilities and back into the community. Unfortunately, few formerly incarcerated individuals are able to access MOUD upon community reentry, remaining at high risk of overdose and rearrest. Community pharmacists are a promising resource for providing injectable naltrexone to formerly incarcerated individuals using this treatment option, but are underutilized during reentry planning and by formerly incarcerated individuals upon release. This is due, in large part, to several barriers that exist across the socioecological scale. Accordingly, this study utilized a participatory design process to inform an intervention that address these barriers and improves access to community pharmacist-provided injectable naltrexone for formerly incarcerated individuals upon community reentry. Three iterative focus groups were conducted with five community pharmacists who have experience providing injectable naltrexone and treating formerly incarcerated patients. The goals of each focus group were to: 1) discuss perceptions of existing barriers and prioritize barriers to be addressed, 2) discuss and rank potential interventions to address the prioritized barriers, and 3) discuss components and anticipated challenges related to the prioritized intervention. Focus groups were analyzed via deductive content analysis using a priori categories. Based on discussions of perceived impact and feasibility, the participants prioritized two barriers to be addressed: lack of awareness of community pharmacist-provided injectable naltrexone services and lack of interagency collaboration among primary care clinics, community pharmacies, and correctional facilities. The final intervention included pharmacist-led educational meetings with correctional providers and reentry staff. Several intervention components and anticipated challenges were also identified. Next steps include developing, implementing, and evaluating the efficacy of the intervention on improving access to community pharmacist-provided injectable naltrexone for formerly incarcerated individuals.

## Introduction

Opioid use disorder (OUD) is defined as a problematic pattern of prescription or illicit opioid use, often leading to serious health and social consequences, including overdoses.^1-2^ In Wisconsin, OUD has become a prevalent public health problem. From 1999 to 2019, there was a 900% increase in opioid overdose deaths.^3^ Notably, OUD is major problem among those impacted by the criminal justice system. From 2013 to 2019, the Wisconsin Department of Corrections reported 1,691 opioid-related hospitalizations among those admitted to probation and 754 opioid-related hospitalizations among those released from prison.^4^

Medications for opioid use disorder (MOUD), which includes long-acting injectable naltrexone, are a critical component in treating OUD.^5^ Due to the high prevalence of OUD among those impacted by the criminal justice system, access to MOUD for these individuals is crucial. While continuation or initiation of MOUD within jails and prisons can still be improved, availability has expanded over the last decade.^6-11^ However, access to MOUD for individuals transitioning out of correctional facilities and back into their communities remains highly limited. For example, in Wisconsin, less than half of jails provide community linkage to MOUD for individuals reentering the community.^7^

Access to MOUD for formerly incarcerated individuals is especially crucial during community reentry. The first few days after release from incarceration present the greatest risk of overdose, as tolerance to opioid is lost while in jail or prison.^12^ Formerly incarcerated individuals receiving MOUD are 85% less likely to die due to drug overdose in the first month after release and have a 32% lower risk of rearrest.^13^ Yet, because so many formerly incarcerated individuals do not have access to MOUD during this time, they remain at a 40-fold greater likelihood of overdose following release compared to the general population.^14^ Additionally, formerly incarcerated individuals account for up to 50% of overdose deaths in certain regions of the country.^15-16^

There is a clear need to increase access to MOUD for formerly incarcerated individuals during reentry. In Wisconsin, a potential resource that may help improve access is community pharmacists. Since 2019, community pharmacists in Wisconsin have the authority to dispense and administer naltrexone injections, a treatment option that shows many benefits and is widely accepted among justice-impacted individuals.^17-18^ However, research shows that there are several barriers to community pharmacist-provided injectable naltrexone for formerly incarcerated individuals, which exist across the socioecological scale.^19^ Overall, it is important that additional work be done to address these barriers.

Accordingly, the goal of this study was to utilize a participatory design process to inform an intervention that reduces the existing barriers and improves access to community pharmacist-provided injectable naltrexone for formerly incarcerated individuals during community reentry in Wisconsin. Participatory design has shown to be beneficial for the design of interventions in complex work systems, including community pharmacies.^20^ Three iterative focus groups were conducted with community pharmacists who had experience providing injectable naltrexone for formerly incarcerated individuals. The focus groups were used to: 1) discuss perceptions of existing barriers and prioritize barriers to be addressed, 2) discuss and rank potential interventions to address the prioritized barriers, and 3) discuss components and anticipated challenges related to the prioritized intervention.

## Methods

### Participants and sampling

Participants were recruited for semi-structured focus groups between March 2024 and April 2024. Study participants included community pharmacists with experience administering naltrexone injections to formerly incarcerated patients. All participants were 18 years of age or older, able to speak and understand English, and residing in Wisconsin. The lead researcher (JC) had established connections to several community pharmacies across Wisconsin and leveraged these connections to identify and recruit participants. Initial recruitment was limited, so snowball sampling was used to identify and recruit additional participants who fit the inclusion criteria. In total, five community pharmacists were recruited. None of the pharmacists worked for the same organization. This study was deemed exempt by the University of Wisconsin-Madison Institutional Review Board (application 2024-0354).

### Procedures

All potential participants were informed of the study and invited to participate via email. Once participants committed to the study, the lead researcher collected availability, and the focus groups were scheduled. An information sheet was then emailed to all participants. The information sheet was reviewed by the lead researcher on the call prior to the start of the first focus group, after which verbal consent to participate was obtained. The lead researcher emphasized that there was no obligation to participate, and participation was voluntary and could be stopped at any time. All focus groups were conducted via Zoom by the lead researcher, who had previous experience conducting semi-structured focus groups. Focus groups were audio recorded to help facilitate transcription and took 1.5 - 2 hours each. After the focus groups, participants were sent a five-minute demographic survey. Participants were compensated with a $100 gift card for each focus group they participated in (up to $300 total).

The researcher conducted three semi-structured focus groups. The focus groups were iterative, with each focus group building off the previous one. Each focus group had specific goals, as outlined in Table 1, that aligned with the first three steps in the participatory design process.^20^ For these goals, impact was defined by which barriers could create the largest improvements if addressed and which interventions would be the most impactful at addressing the prioritized barriers. Feasibility was defined by which barriers could be practically addressed and which interventions could be practically implemented.

**Table 1.** Focus group goals.

| Focus Group # | Goals |
| --- | --- |
| 1 | • Discuss perceptions of existing barriers<br>• Prioritize barriers based on perceived impact and feasibility |
| 2 | • Discuss potential interventions to address the prioritized barriers<br>• Rank intervention ideas based on perceived impact and feasibility |
| 3 | • Discuss intervention components<br>• Discuss anticipated challenges related to the intervention |

The same group of community pharmacists participated in all three focus groups. For each focus group, the lead researcher developed a guide to help prompt the discussions. Additionally, the lead researcher utilized a digital whiteboard from Mural, an online collaboration tool, to take notes on the meetings.^21^ The digital whiteboard was shared in real time during the focus groups so that participants could visually track the conversations and make better connections between ideas. At the end of each focus group, participants were given the opportunity to share any thoughts or ideas that had not been addressed by the questions in the guide. All focus groups took place from April 2024 to May 2024.

### Data coding and analysis

The focus groups were transcribed verbatim, de-identified and verified for accuracy. All participants were assigned an ID number. Transcripts were entered into NVivo, a qualitative data software package (released in March 2020).^22^ As outlined by Elo & Kyngäs, two independent coders then performed deductive content analysis to place data into a priori categories.^23^ The categories were based on the questions from the focus group guide and are outlined in Table 2.

**Table 2.**
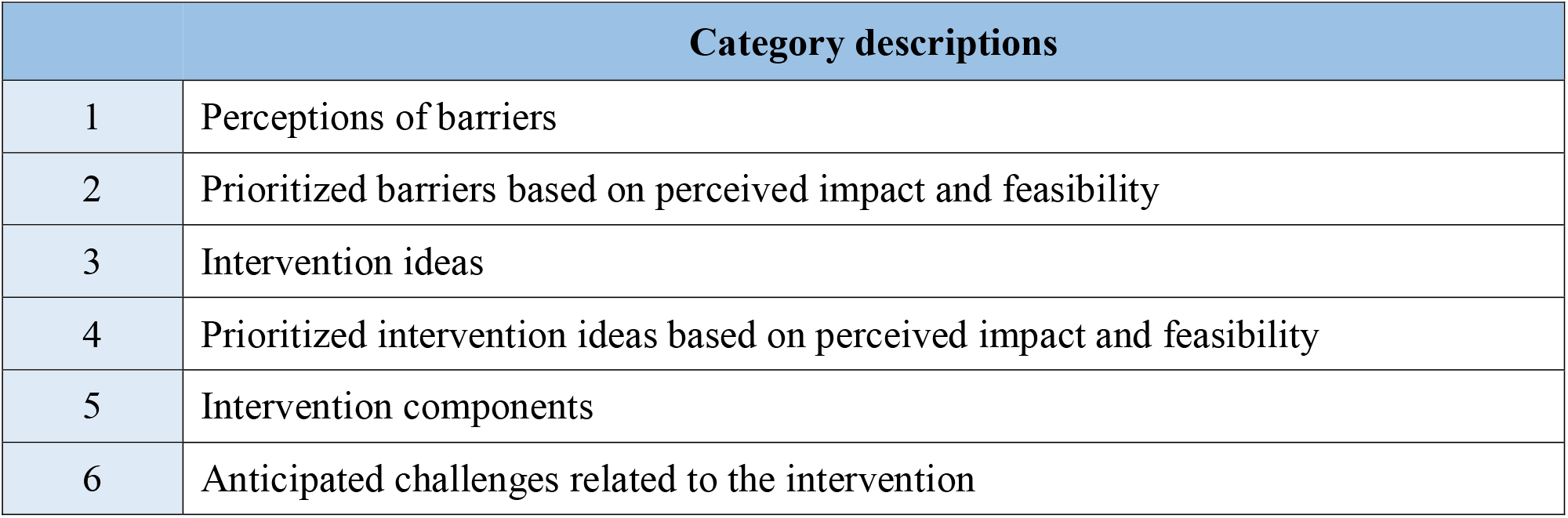
A priori categories for deductive content analysis of focus groups.

This analysis process was used, as participants occasionally discussed information that was relevant to questions from a different focus group. For example, although focus group 3 included questions related to anticipated challenges, some participants mentioned challenges as they prioritized intervention ideas in focus group 2. As a result, it was more effective to code all focus groups across the same categories.

After the coding process, the lead researcher summarized the data within each category. The research team met to discuss the summaries, as well address any ambiguities or issues related to coding. Finally, representative quotes were selected to support the results. Overall, the four-dimension criteria of qualitative research were used to guide the data coding and analysis process.^24^

## Results

In total, five community pharmacists participated in all three focus groups. Participant demographics are outlined in Table 3. Results from the focus group are described below and separated based on the focus group number and data categories utilized during analysis. Overall, many of the participants expressed similar thoughts throughout the focus groups, especially regarding their perceptions of the existing barriers to community pharmacist-provided injectable naltrexone for formerly incarcerated individuals. Any variations or nuances between participants are discussed where applicable.

**Table 3.**
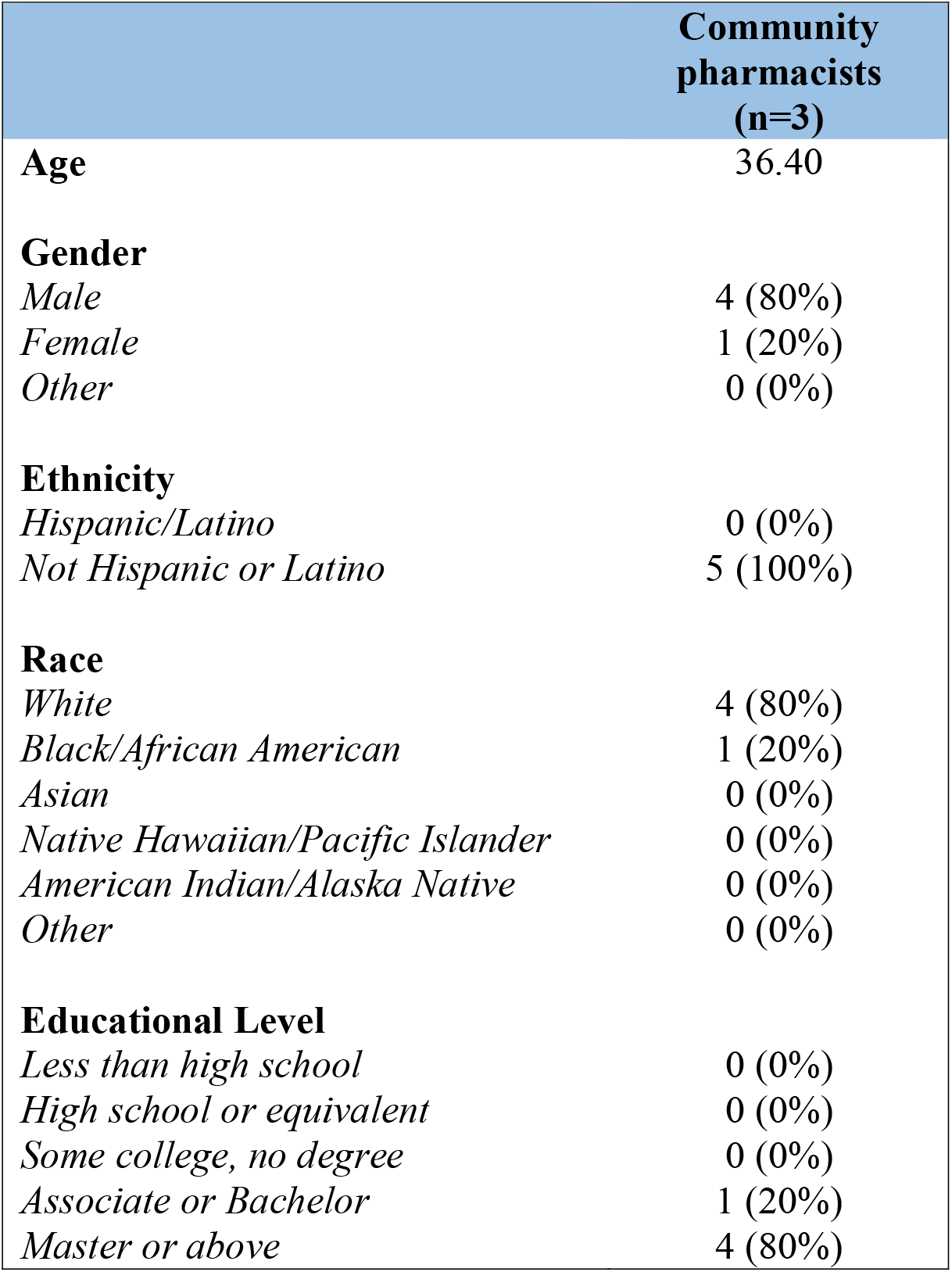
Participant demographics.

|  | **Community pharmacists**<br>**(n=3)** |
| --- | --- |
| **Age** | 36.40 |
| **Gender** |  |
| *Male* | 4 (80%) |
| *Female* | 1 (20%) |
| *Other* | 0 (0%) |
| **Ethnicity** |  |
| *Hispanic/Latino* | 0 (0%) |
| *Not Hispanic or Latino* | 5 (100%) |
| **Race** |  |
| *White* | 4 (80%) |
| *Black/African American* | 1 (20%) |
| *Asian* | 0 (0%) |
| *Native Hawaiian/Pacific Islander* | 0 (0%) |
| *American Indian/Alaska Native* | 0 (0%) |
| *Other* | 0 (0%) |
| **Educational Level** |  |
| *Less than high school* | 0 (0%) |
| *High school or equivalent* | 0 (0%) |
| *Some college, no degree* | 0 (0%) |
| *Associate or Bachelor* | 1 (20%) |
| *Master or above* | 4 (80%) |

### Focus Group 1

#### Perceptions of barriers

The participants were first presented with the prevalent barriers related to community pharmacist-provided injectable naltrexone for formerly incarcerated individuals. These included: lack of reliable transportation, lack of insurance, lack of interagency collaboration between primary care clinics, community pharmacies, and correctional facilities, lack of awareness of community pharmacist-provided injectable naltrexone services, inability of pharmacists to provide additional OUD services, stigma, drug cost, and lack of available prescribers and injectors.^19^ Participants were first asked about their initial perceptions of these barriers, or if any came as a surprise. Overall, the participants stated that upon initial review, each of the barriers made sense and aligned with their perception of the current situation. One participant stated, “From my perspective, these all make sense. Especially knowing that not a lot of community pharmacies offer injectable naltrexone, at least to my knowledge,” (RPh1). The rest of the community pharmacists had similar reactions and, accordingly, none of them pointed to barriers that were particularly surprising.

Additionally, the participants noted that many of the barriers overlapped. One pharmacist pointed out that, “All of them line up appropriately. Especially the collaboration with primary care and correctional facilities, which kind of goes hand in hand with them now knowing that community pharmacies are able to provide this service,” (Rh4). Another pharmacist noted that the inability of pharmacists to provide additional OUD services directly relates to the lack of available injection sites. Only one barrier received minimal pushback, as one participant noted that stigma might not be a major barrier at every pharmacy, depending on whether or not the pharmacy has the ability to offer a private room for injections. If patients are aware that they can receive treatment privately, they may be less concerned with experiencing stigma.

#### Prioritized of barriers based on perceived impact and feasibility

To help prioritize which barriers should be targeted by a potential intervention, the participants were asked to think about which barriers, if addressed, could create the biggest improvement in access to community pharmacist-provided injectable naltrexone for formerly incarcerated individuals. In other words, how impactful addressing a particular barrier would be. In terms of impact, the pharmacists focused on five of the eight barriers. These five barriers, as well as representative quotes, are outlined in Table 4.

**Table 4.** Prioritized barriers based on perceived impact.

| **Barrier** | **Representative quotes** |
| --- | --- |
| Lack of awareness | “Increasing awareness of services – from the patient all the way down to the person that is leading them out of the [correctional facility]. If they know that [community pharmacists] are involved in giving these injections, that’s huge. Increasing awareness, especially through increasing marketing, would be so much more impactful than connecting with [patients] by making a bunch of phone calls.” - RPh2 |
| Lack of interagency collaboration | “The collaboration. Because it is so important to make sure that the provider, the [reentry staff], and the pharmacist are on the same page.” - RPh1 |
| Stigma | “I would add stigma, especially from [the patient’s] perspective. Making sure that they feel welcome and have the ability to access medications without judgement – both from a medical history and social history perspective – that is going to lean a lot into improving access.” - RPh3 |
| Inability of pharmacists to provide additional OUD services | “I think providing additional services, specifically the drug testing. If we can get down, I think, from the perspective of the pharmacy world, would help a lot.” - RPh4 |
| Drug cost | “Money makes the world run. So, not necessarily lowering the cost of the drug, but at least showing a cost-benefit, and that we are profiting off of this could really help.” - RPh2 |

The participants were then asked to think about which barriers could be most feasibility addressed. The conversation around feasibility focused on three of the eight barriers. These barriers, as well as representative quotes, are outlined in Table 5.

**Table 5.** Prioritized barriers based on perceived feasibility.

| **Barrier** | **Representative quotes** |
| --- | --- |
| Lack of awareness | “Increasing awareness is [feasible]. It can happen through simple discussion with staff or with a provider. Like, ‘Hey, just to you know, we provide these services.’” - RPh1 |
| Lack of interagency collaboration | “Community pharmacists have had a successful history of creating collaborations with physicians, with practitioners for other services. So, I think that would a feasible option here.” - Rh4 |
| Lack of available prescribers and injectors | “I think increasing the number of community pharmacists who provide injections, just by getting them trained. Even if it isn’t naltrexone injections right away, but just another type of injection to get them comfortable.” – RPh1 |

Lastly, based on the discussions surrounding perceived impact and feasibility, the pharmacists were asked to select one or two of the barriers that they would target with an intervention. Unanimously, the participants selected lack of awareness and/or lack of interagency collaboration. These decisions were largely based on the fact that these barriers were perceived to be both feasible and impactful if addressed. As a result, these two barriers were used as the basis for focus group 2. An image of the Mural digital whiteboard and notes from focus group 1 are included in Figure 1.

**Figure 1.**
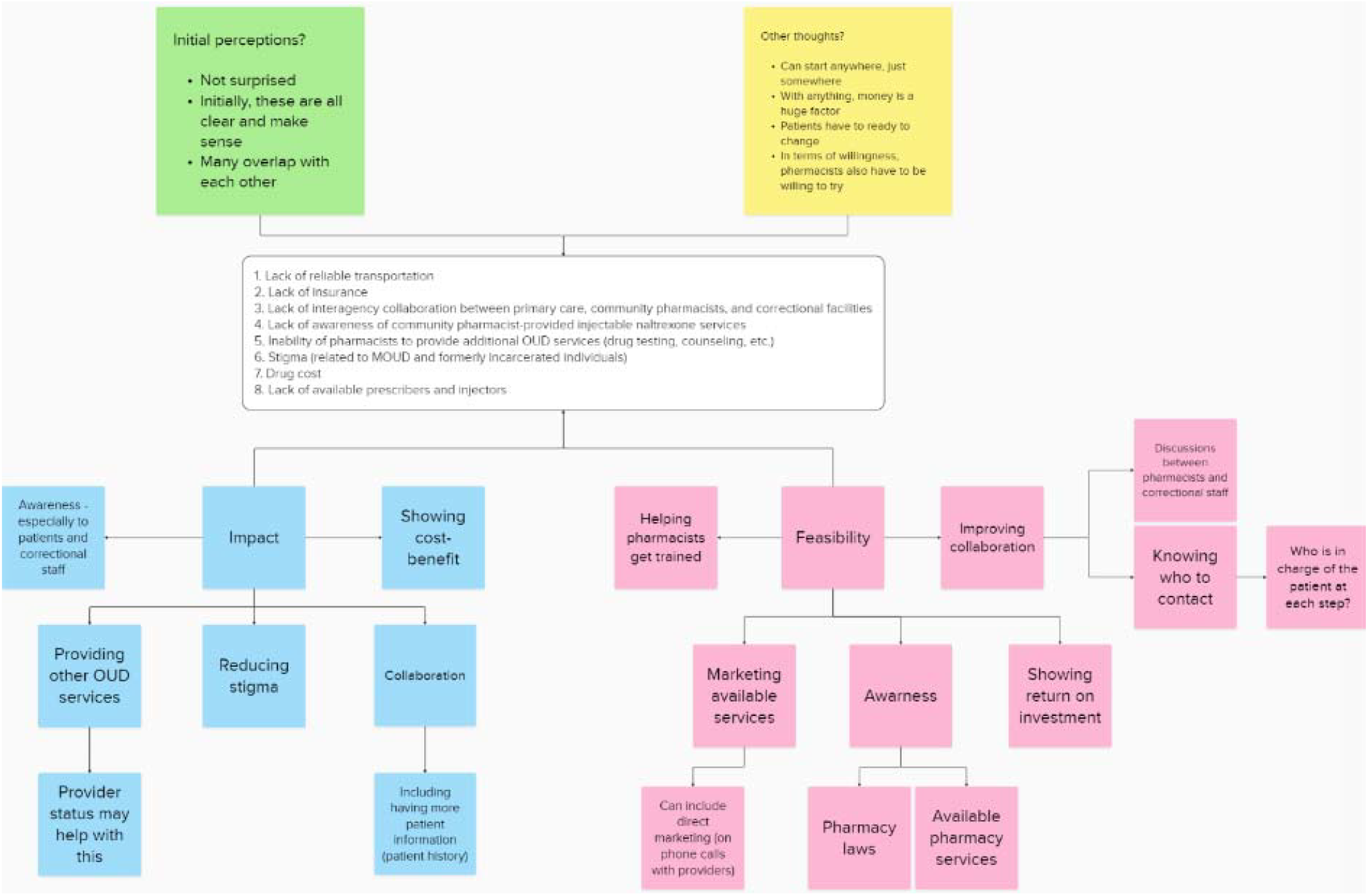
Mural digital whiteboard from focus group 1.

### Focus Group 2

#### Intervention ideas

Based on the results of focus group 1, the participants were instructed to focus their discussion on two barriers of interest: 1) lack of awareness of community pharmacist-provided injectable naltrexone services and 2) lack of interagency collaboration between primary care clinics, community pharmacies, and correctional facilities. First, the participants were asked to brainstorm interventions that could address at least one of these barriers, and several ideas were shared. These ideas are outlined in Table 6 and supplemented by representative quotes.

**Table 6.** Intervention ideas.

| Idea | Representative quotes |
| --- | --- |
| Development of a recovery clinic | “An opportunity could be, like, building out a recovery clinic where there are particular mental health providers, nurses, that can team up with community pharmacists that can dispense and provide the injection. I just think it might help to have, you know, streamlined services to specific clinics that might already have a rapport built up for opioid use disorder and have it be sent to those particular pharmacies that are providing those services.” - RPh5 |
| Adding community pharmacists to existing online resources | “I think a good first step is to get in line with the Vivitrol website. I think it’s quite literally just vivitrol.com and you can find a provider. I think [providers] have to manually add themselves to that, so that’d be a really good starting point...getting [community pharmacists] as providers on that website would be a good first step.” - RPh1 |
| Development of an informational website | “Maybe, you know, we do have a website for the case managers and those who are going to be helping connect the dots that states here are all the different, you know, pharmacies that are going to be giving long-acting injections in particular. You know, this is the insurance that they take...I think there could be a website specifically for opioid use disorder.” - RPh1 |
| Development of an informational pamphlet | “We could have a little pamphlet that says, you know, it kind of goes through, you know, what opioids are. But on the front, there’s a little QR code...that brings them to [injectable naltrexone] near me or something along those lines. Or where I can find a pharmacy that carries [injectable naltrexone] and accepts certain insurance.” - RhP4 |
| Development of a central repository document | “I think there needs to be some type of central repository document. We already know that there are some places offering these services. But being able to see where these services are for, again, the staff that would help with reentry and ultimately that can get them connected to a pharmacy near that person’s home, that would be very helpful.” - RPh5 |
| Pharmacist-led educational meetings with correctional staff | “Maybe setting up meetings with some of the [correctional staff] and just letting them know that this is something that we offer...I think it’s building that rapport and just opening the door and saying, ‘Hey, this is something that we’re offering at the pharmacy.’” - RPh4<br>“Meetings are a great starting point. And I think education really needs to start [within corrections]. If you’re looking at this specific patient population of how they are falling through the cracks and how they are winding up back behind bars, I think that is where it starts.” - RPh1 |
|  | “I feel like we need to give education even further downstream. So, like, the prisons and facilities where those people who are giving the injections or the providers who did prescribe that injection in the prison can be able to then find the resources to connect them to the particular [community pharmacies] they could be. I almost feel like that’s where it should start at first.” - RPh5 |

#### Prioritized intervention ideas based on perceived impact and feasibility

Participants were asked to consider which intervention, if implemented, could create the biggest improvement in access to community pharmacist-provided injectable naltrexone for formerly incarcerated individuals. They were also asked to consider which intervention would be most feasible to implement as a starting point. Not only did several community pharmacists identify community pharmacist-led educational meetings with correctional staff as a potential solution, but this intervention was almost immediately prioritized by the participants. In thinking about impact, one participant stated, “Yeah, if there’s anything coming out of this, it’s education so that [correctional staff] understand that community pharmacies offer [injectable naltrexone] services and understand the steps to use them. That education needs to happen. It would be the best thing to come from this,” (RPh3). The rest of the participants agreed with this statement and added that educating correctional staff on available community pharmacist-provided injectable naltrexone services could create a significant impact on connecting formerly incarcerated individuals to these treatments. Additionally, the participants unanimously agreed that educational meetings would not only be a feasible option, but provide the best balance between impact and feasibility.

The participants mentioned several other reasons that pharmacist-led educational meetings with correctional staff should be a prioritized intervention. First, a few of the pharmacists stated that it is important to start at the source of the problem. Since formerly incarcerated individuals are reentering the community from correctional facilities, an intervention should be targeted at those who are involved in reentry at that point in time. Second, the participants explained that these meetings could accomplish several tasks. For example, the meetings could not only be used to increase awareness of community pharmacist-provided injectable naltrexone, but could also help educate correctional staff on utilizing prescriptions and what patient information is required by community pharmacists, be used as an outlet to share existing resources, and allow pharmacists and correctional staff to establish points-of-contact. Importantly, the participants mentioned that these meetings could help address both of the prioritized barriers by increasing awareness and, in the long-term, increasing collaboration among community pharmacists and correctional staff.

In terms of the other interventions that were suggested, a few community pharmacists noted that while some were good ideas, they wouldn’t be as impactful at improving access to community pharmacist-provided injectable naltrexone for formerly incarcerated individuals. For example, adding community pharmacists as providers to online resources may be helpful, but it would still require correctional staff and/or formerly incarcerated individuals to be aware of these resources and leverage the information on their own. Similarly, a few pharmacists noted that some of the intervention ideas would not be as feasible. Notably, while developing a recovery clinic could be very beneficial as a long-term goal, the participants mentioned that this would be difficult to implement as a first step. Based on all of these thoughts, pharmacist-led educational meetings with correctional staff was selected as the prioritized intervention. An image of the Mural digital whiteboard and notes from focus group 2 are included in Figure 2.

**Figure 2.**
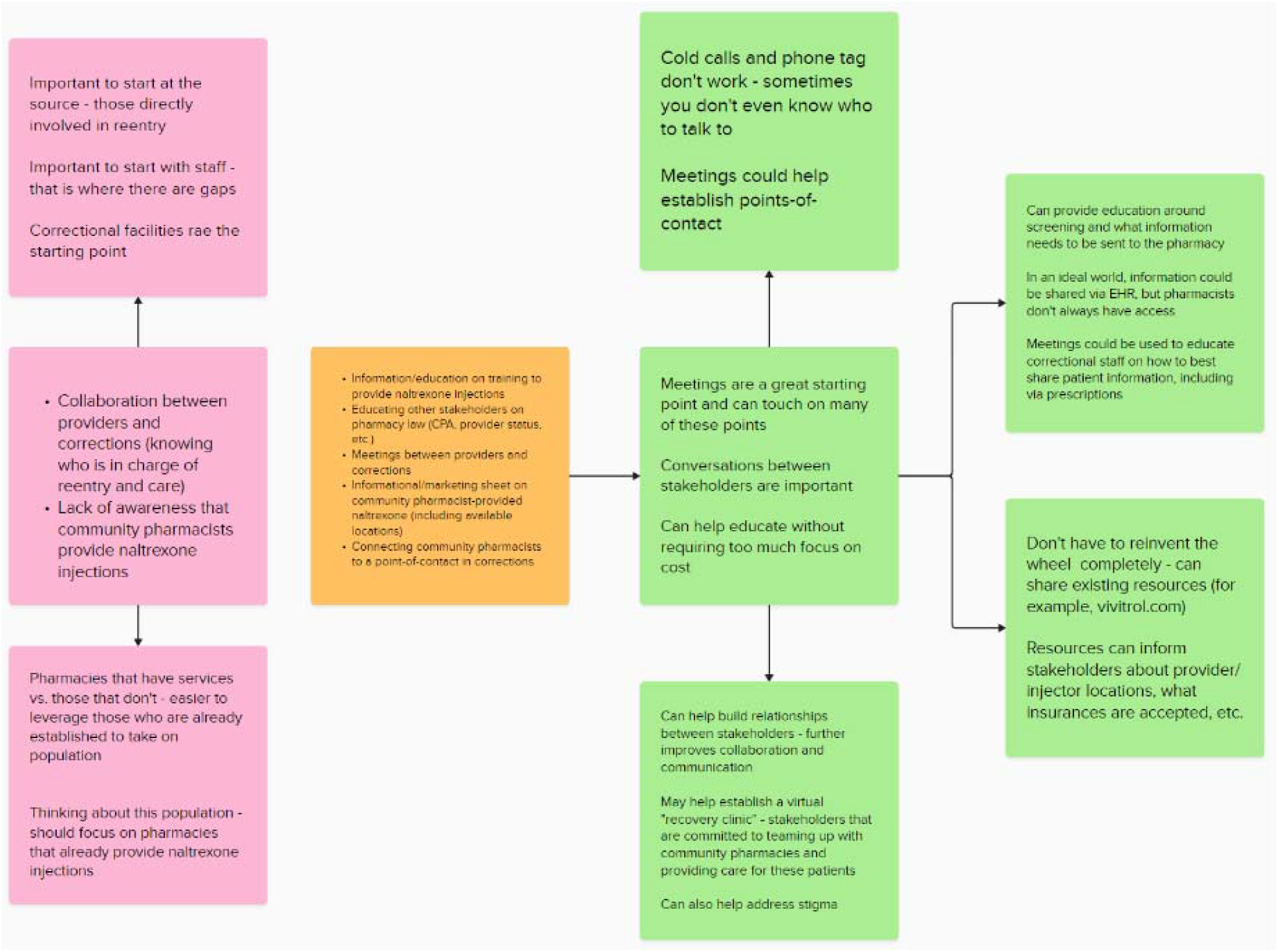
Mural digital whiteboard from focus group 2.

### Focus Group 3

#### Intervention components

Based on focus group 2, the participants selected pharmacist-led educational meetings with correctional staff as the intervention of interest. In order better conceptualize and inform the development of this intervention, the participants were asked to identify components that should be included in the educational meetings, in addition to letting correctional staff know that community pharmacists are able to provide injectable naltrexone. Additional components, as well as representative quotes, are outlined in Table 7.

**Table 7.** Intervention components.

| Component | Representative quote |
| --- | --- |
| Sharing existing resources | “I’m a big fan of not reinventing the wheel. It would be helpful not to reinvent the wheel on everything. So, maybe some of these resources, like [vivitrol.com] could be shared during the meetings.” – RPh3 |
| Educating on required patient information | “I think it’s important to educate on the things that are required at the pharmacy end. I think it needs to be known really by anyone who is involved with injections or reentry. Things like a diagnosis code, history of using [injectable naltrexone], date of last injection, if they’ve tried oral naltrexone...” - RPh4 |
| Educating on utilizing prescriptions to provide patient information | “In an ideal world, we would love to be connected with EHR. But I even think on the prescription itself, there’s an area that says ‘Pharmacy Notes’ that you can actually input criteria, like, you know, ‘Yes, they’re a candidate, this is the last time they took the drug, they have taken this medication before...’ I think that would be helpful for the [correctional staff] to know, so [the pharmacist] can have some information if they’re not connected to the HER.” - RPh5 |
| Establishing points-of-contact | “Sometimes you’re trying to call and, you know, schedule appointments...or if [a formerly incarcerated individual] missed their appointment to try and call and get them back in for, you know, a reschedule...quite often their phone number changes, or their voicemail box has not been set up. Or you don’t even have an actual address on file because they’re kind of in that transitional stage where they are moving around and kind of getting reestablished. So, using [the meetings] to establish points-of-contact or contact info for social workers or case management can be huge.” - RPh1 |
| Emphasizing cost-benefit | “Make sure you mention any kind of monetary incentive for them because it’s expensive to have somebody in jail and go back to jail. Injectable naltrexone is also expensive, but I could only assume that having them on monthly injection as opposed to having them in jail for another month at minimal...there’s a benefit of savings right there.” - RPh4 |
| Educating on importance of enrolling individuals in insurance | “Maybe just insurance considerations. If we’re talking about someone transitioning from a correctional facility to home or wherever, insurance factors into that. So, maybe including insurance considerations. And making sure that, like, the reentry staff knows they need to help these individuals kind of access insurance first before anything.” - RPh1 |

Overall, the participants recognized that the focus on the meetings should be educating correctional providers and reentry staff on the ability of pharmacists to provide naltrexone injections for individuals transitioning out of correctional facilities and back into the community, as well as the fact that community pharmacists are accessible providers. However, the participants also agreed that the additional components outlined above could make the meetings more impactful without adding a significant amount of additional work.

The participants were also asked about how the educational meetings would best be delivered. Overall, two main considerations emerged. First, the pharmacists all agreed that the meetings should be held in-person. One participant said, “I think in-person meetings are always going to be a lot easier and more people are able to digest more information,” (RPh3). Another stated, “I vote in-person. I think you can build more relationships that way and you can, you know, answer questions that might come up a little bit easier if you’re in person. Things you might not have thought of when you were developing a web module or handout,” (RPh4). A third added, “Yeah, I second or third in-person. For me, I think it’s like, you know, building those relationships with people and kind of being able to express how emotionally invested you are as opposed to trying to imitate that via a webinar…I think having somebody in-person that can really say, ‘I’ve seen this change people’s lives.’ Simple as that,” (RPh1).

In addition to pushing for in-person meetings, the participants agreed that the educational meetings should be led by pharmacists who have experience providing naltrexone injections and working with formerly incarcerated patients. “I think it’s definitely easier for a pharmacist that’s already established [these services] to kind of take the lead on this,” stated one pharmacist (RPh1). Another echoed this thought and added, “And if you have somebody from a community pharmacy that is already offering this, you automatically make that connection. So, a really good strength of having [pharmacists with experience] lead is that you’re creating those connections right away for those [correctional] facilities,” (RPh4).

Lastly, the focus group participants were asked if there were any other stakeholders that should be included or invited to the educational meetings. Overall, the participants agreed that correctional staff (providers and reentry coordinators) should be the center of the meetings.

However, there were a few additional stakeholders that the participants thought could either improve the meetings or benefit from the information shared during the meeting. One participant said that drug representatives could support the pharmacists in educating correctional staff. “One [stakeholder] that comes to mind is drug reps…They have the time and they’re getting paid, and they can help with the educational piece,” (RPh1). Another participant added, “I think what we’re missing here is not involving social work or case management in the discussion. They really help bridge, so I would actually add having them involved in the discussion when you are having these in-person meetings,” (RPh5). Lastly, one pharmacist said that it would be beneficial to involved governmental officials. They said, “I would say include someone as high up in the government for the state as you can, too. Because if you can get, like, governor’s office on board or whoever the state overseer for correctional facilities is, like, and we make it a state priority, I think you’ll get a lot more buy-in from the facilities themselves,” (RPh4).

#### Anticipated challenges related to the intervention

Finally, the participants were asked to identify any challenges that they anticipate with developing and/or implementing community pharmacist-led educational meetings with correctional staff. Overall, three main challenges were identified. First, one participant expressed concerns with overuse of injectable naltrexone among formerly incarcerated individuals. They stated, “So, with something like injectable naltrexone, the last thing I would want to happen is that they recognize that they can give injectable naltrexone and they start slapping it on every person that leaves that has opioid use disorder. And then these folks go back and use right away afterwards, and we have a lot more complications,” (RPh4). Another participant expressed a similar concern, saying, “Providing education solely on naltrexone, on the injectable form, could lead – especially if they don’t have a healthcare background – it could lead to some institutions just automatically jumping to injectable in patients that it’s not idea for, which is a huge risk to that person and could lead to some really poor outcomes for those folks,” (RPh1). Second, one participant said that time might be a challenge or barrier. “I think a second thing is that if we focus on individual education or, like, institution to institution, it’s going to be very time consuming, even if we have the partnerships and everything like that,” (RPh1). Third, some of the pharmacists expressed concerns about who would be able to attend in-person meetings and whether or not those in rural areas would be excluded. “And obviously there’s going to be a lot of places in the rural settings that they’re not able to meet in person, so showing that you’re, like, fully invested in this, that would be very helpful,” (RPh5). An image of the Mural digital whiteboard and notes from focus group 3 are included in Figure 3.

**Figure 3.**
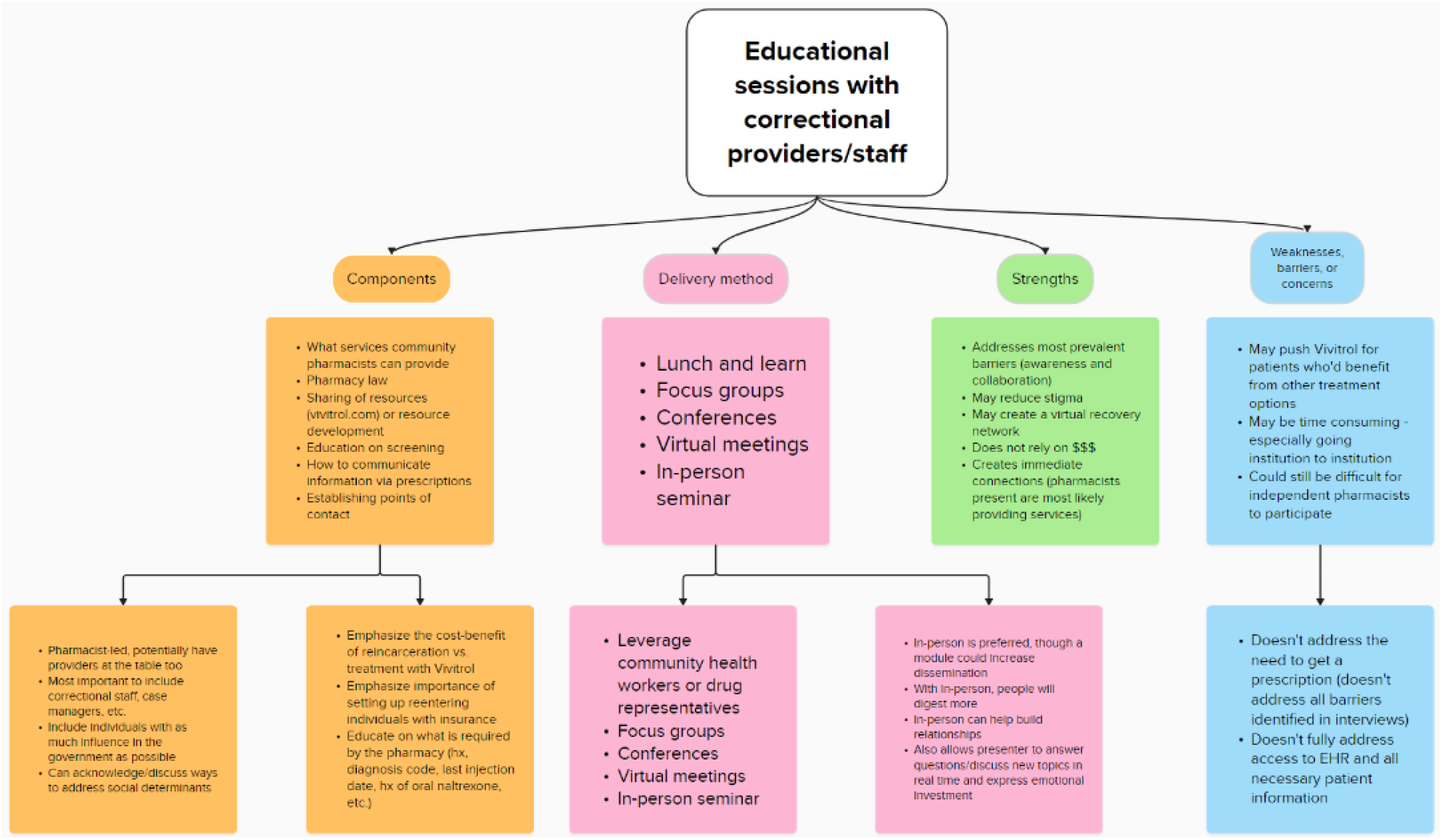
Mural digital whiteboard from focus group 3.

## Discussion

Throughout the focus groups, participants were given the opportunity to discuss and prioritize the barriers related to community pharmacist-provided injectable naltrexone for formerly incarcerated individuals, as well as inform an intervention to address these barriers. Across all three focus groups, there was a high level of agreement among the participants. In terms of focus group 1, the pharmacists had similar perceptions of the existing prevalent barriers, agreeing that the barriers made sense related to their existing knowledge. Overall, this is not surprising, given that the participants had experience providing injectable naltrexone for formerly incarcerated patients and had likely experienced many of the barriers first-hand.

During the focus groups, the participants unanimously agreed that the intervention should be targeted upstream with correctional staff. This is important, as we know that the first several days after community reentry present the greatest risk to formerly incarcerated individuals with OUD. As a result, developing an intervention that targets corrections can help connect formerly incarcerated individuals to community pharmacist-provided injectable naltrexone as soon as they reenter. For example, it may also be beneficial to implement an intervention that increases the number community pharmacies providing injectable naltrexone or helps community pharmacies provide additional OUD services. However, without awareness of these services by correctional staff and/or collaboration between corrections and community pharmacies, formerly incarcerated individuals may still be left to find and access these services on their own.

Notably, the intervention that was informed by the focus group participants can help address both of the prioritized barriers (lack of awareness and lack of collaboration), which may be especially impactful. Not only that, but the specific intervention components added by the focus group participants can help address other barriers that were previously identified, making the meetings a multipurpose intervention.^19^ For example, the participants stated that during the educational meetings, the pharmacists leading the meetings should stress the importance of enrolling individuals in insurance before they reenter the community. This can push reentry staff to make enrollment a priority, helping to increase insurance access for formerly incarcerated individuals before they are back in the community. Notably, in order to address the opioid epidemic in Wisconsin, the Department of Corrections (DOC) previously created trainings to educate staff about the three MOUD options.^7^ Ultimately, the intervention informed by the focus groups aligns with other opioid-focused interventions that have been implemented in correctional settings across Wisconsin.

In thinking about how to set up the educational meetings, all of the focus group participants agreed that the meetings should be led by community pharmacists with experience providing injectable naltrexone and working with formerly incarcerated individuals. However, several of the participants identified other stakeholders that could benefit from the content shared during the meetings. These included drug representatives, social workers and/or case managers, and governmental officials. Including (or at least inviting) these professionals to the educational meetings could be beneficial, as it not only incorporates other perspectives, but can further improve awareness of community pharmacist-provided services and foster even more collaborative relationships.

In addition to these professionals, one stakeholder that was not mentioned was community health workers. Community health workers are individuals from the community who form relationships with individual patients and assist them in accessing health care and health-related resources.^25^

Importantly, community health workers can help patients overcome barriers related to the social determinants of health. Previous work has shown the benefits of collaborations between community health workers and pharmacists in improving patient outcomes.^25-26^ As a result, integrating community health workers into this intervention could also prove to be beneficial, especially considering many of the obstacles that formerly incarcerated individuals face in accessing care. Furthermore, it may be beneficial to include community pharmacists who don’t have experience providing injectable naltrexone and/or working with formerly incarcerated individuals. While the focus group participants agreed that those with these experiences should lead the meetings, inviting other pharmacists offers them the chance to learn more about integrating injectable naltrexone services into their practice and/or the impact they can make by connecting with and treating formerly incarcerated patients. This could also help address the lack of injection sites across Wisconsin.

As demonstrated by the third focus group, the intervention is not without potential challenges. However, there are strategies that could help eliminate or at least mitigate some of these barriers. For example, the participants expressed concerns about only educating on injectable naltrexone, stating that they wouldn’t want this option used for every formerly incarcerated individual with OUD. To prevent this problem, the pharmacists leading these meetings could educate on which patients benefit the most from injectable naltrexone, discuss how to screen for these patients, and emphasize that injectable naltrexone is not the best treatment option for all individuals with OUD. They could also briefly discuss the other forms of MOUD that exist and highlight some resources that provide guidance on accessing these options if necessary. Additionally, the participants said that implementing educational meetings could be time consuming, and some expressed concerns that correctional staff in rural areas would be excluded. One way to overcome these challenges is by coordinating meetings that involve correctional staff across a certain region of Wisconsin. By scheduling these meetings in advance and utilizing central locations, those residing in rural areas may have an easier time attending. At minimum, recordings could be sent to those who are unable to attend in-person meetings.

Next steps should include the development and implementation of the educational intervention. It is likely that the final structure and/or content of the intervention material will have to go through several iterations. Utilizing a community-engaged process can help ensure that the intervention is as meaningful and effective as possible. The intervention should also be assessed for acceptability, appropriateness, and feasibility among a larger group of community pharmacists and correctional staff. Additionally, the researchers should connect with community pharmacists and correctional staff who would be willing to participate in pilot trials of the educational meetings, specifically those in areas where community pharmacist-provided injectable naltrexone is already available. These trials could be used to demonstrate the efficacy of the intervention on increasing correctional staff knowledge of community pharmacist-provided injectable naltrexone services, collaboration between correctional facilities and community pharmacies, and the use of community pharmacist-provide injectable naltrexone by formerly incarcerated individuals upon reentry. Showing potential efficacy can support the scale-out of the intervention to other areas, including areas outside of Wisconsin.

### Limitations

There are a few study limitations that should be noted. For starters, because the study used convenience and snowball sampling to recruit community pharmacists, it is possible that bias was introduced. Additionally, the community pharmacists included in this study were from several counties in Wisconsin, including urban and rural areas. However, since pharmacists from every area couldn’t be included, it is possible that the results do not represent the opinions and ideas of all pharmacists across Wisconsin. The results may also not be generalizable to areas outside of Wisconsin. The study was also limited to community pharmacists who had experience providing injectable naltrexone for formerly incarcerated individuals. This may have influenced their decisions regarding the barriers and potential interventions, especially considering the resulting intervention was an education-based intervention. Ultimately, the inclusion criteria and nature of the selected intervention likely limit next steps to areas that have existing community pharmacist-provided injectable naltrexone services. It is also likely that the pharmacists who choice to participate in this study were more open to collaboration, potentially influencing their perceptions and limiting generalizability. Finally, the participants were predominantly male, white, and did not identify as Hispanic or Latino, resulting in a homogenous sample. Despite these limitations, this study was intended to be exploratory in nature, and additional work can help ensure the transferability of results.

## Conclusion

Overall, the results of this study can help increase awareness of community pharmacist-provided injectable naltrexone services among correctional providers and reentry staff in Wisconsin.

Long-term, this can help increase collaborations between correctional facilities and community pharmacies and, as a result, the use of community pharmacist-provided injectable naltrexone by formerly incarcerated individuals upon reentry. Increased access to injectable naltrexone can help improve several health and social outcomes for this patient population. Importantly, MOUD access can help formerly incarcerated individuals avoid the cycle of rearrest and reincarceration and be able to thrive in society.

## Data Availability

All data produced in the present study are available upon reasonable request to the authors.

## Acknowledgements

The authors would like to thank James H. Ford II and Olayinka O. Shiyanbola for their contributions in conceptualizing this manuscript.

